# Effectiveness of iso-inertial resistance training on eccentric and concentric power, physical performance, and risk of falls in physically active middle-older adults: a randomised controlled trial

**DOI:** 10.1101/2024.05.13.24307107

**Authors:** Aïda Cadellans-Arróniz, David Blanco, Marc Madruga-Parera, Victor Zárate-Lozano, Flora Dantony, Daniel Romero-Rodríguez

**Affiliations:** Department of Physiotherapy, Universitat Internacional de Catalunya, Barcelona, Spain

**Keywords:** Middle-older adults, iso-inertial training, resistance training, strength training, eccentric power, flywheel

## Abstract

**Objective:** To evaluate the effectiveness of iso-inertial resistance training on eccentric power compared to gravitational training in physically active middle-older adults.

**Methods:** Parallel-group, randomised controlled trial at Espai Esport Wellness Center (Granollers, Spain). Forty-four physically active adults (>57 years of age) were randomised to iso-inertial (n=21) or gravitational (n=23) training groups (R software; 1:1 ratio). Participants had to complete a 6-week training program (2 sessions/week) consisting of three exercises (forward lunge, side lunge, forward lunge with row). Primary outcome: power in the eccentric phase of each exercise evaluated with both iso-inertial and gravitational devices. Secondary outcomes: concentric power, physical performance, risk of falls. Only outcome evaluators were blinded. We used multivariate linear regression models to analyse the effect of interventions.

**Results:** Iso-inertial training showed better eccentric power gains than gravitational training for iso-inertial system evaluation, although the difference was only statistically significant for the side lunge. Forward lunge: between-group difference 3.99 W (95% CI: -3.99 to 11.33, p: 0.28); side lunge: difference 8.50 W (95% CI: 2.13 to 14.87; p: 0.01); forward lunge with row: difference 14.07 W (95% CI: -2.07 -to 30.20, p: 0.09). We observed no differences for the gravitational system evaluation nor for concentric power, physical performance, and risk of falls. The two groups improved remarkably from baseline for all outcomes.

**Conclusions:** Iso-inertial resistance training might lead to better eccentric power gains than gravitational training. Both approaches seem equally effective in improving concentric power and physical performance, and reducing the risk of falls.

**Trial registration:** ClinicalTrials.gov (NCT06160089).

WHAT IS ALREADY KNOWN ON THIS TOPIC

- Physical exercise in middle-older adults is an effective strategy to promote health and improve quality of life.
- Resistance training using iso-inertial devices generates an advantage in hypertrophy, electromyographic activity or balance compared to cable-resistance training in middle-older adults.
- The power at which an action is performed is considered a predictor of functional capacity, as it is associated with the execution of activities of daily living such as climbing stairs, getting up from a chair or walking.

WHAT THIS STUDY ADDS

- The iso-inertial training system improves muscle power in the eccentric phase compared to the gravitational system, although the difference is only statistically significant for the side lunge exercise.
- Iso-inertial and gravitational resistance training are equally effective in improving concentric power, and physical performance and reducing the risk of falls.
- Using iso-inertial devices is recommended to evaluate power in the eccentric phase, as they may capture better the eccentric demands than gravitational devices.

HOW THIS STUDY MIGHT AFFECT RESEARCH, PRACTICE OR POLICY

- Using iso-inertial devices for resistance training in middle-older adults seems a promising way to improve the power in the eccentric phase of an action.
- Improving eccentric power in older adults is crucial due to its transfer to daily life activities.
- Regardless of the training system, clinicians should prescribe resistance training programs to middle-older adults as these programs remarkably increase power and physical performance and reduce the risk of falls.

## Introduction

### Background and objectives

With the population’s progressive increase in life expectancy, attention to ageing has attracted increased interest in recent years. Actions promoting healthy ageing are key to slowing the physiological progressive loss of skeletal muscle mass and function as a person ages. In particular, muscle strength and power are reduced during aging, harming older adults’ functional capacity and quality of life (1–3). Recent research has revealed a significant decrease in muscle strength of 1.5-5% each year from age 50 (4). Muscular power is the maximum force one can generate during a specific movement at a specified velocity. It is considered a predictor of functional capacity, as it is associated with the execution of activities of daily living such as climbing stairs, standing up from a chair, or walking. Previous studies have reported that power diminishes more than strength over time (5). Both strength and power training have been implemented to prevent the risk of falls and improve the balance or walking capacity in older adults (2,3).

Resistance training (RT) is one of the main strategies used to prevent decreased functional capacity and has been demonstrated to be effective in combating age-induced muscle atrophy (sarcopenia), the risk of falls, and fragility (6). It has been observed that compared with RT with concentric-based exercises, RT with eccentric-based exercises leads to greater maximal strength with less muscle activation and lower metabolic cost, and a greater increase in muscle mass (7,8).

The most traditionally used RT method in community health is the gravitational method, in which resistance is opposed through free weights by blocks in cable machines. The gravitational method is considered one of the most functional resistance training approaches (9). One of the main limitations of the gravitational method is that the workload applied in the concentric phase of the lengthening-shortening cycle limits the muscle capacity progression in the eccentric phase. This fact limits the potential of this method for generating the improvements associated with eccentric training.

In contrast, in the iso-inertial training method, the resistance is generated by an iso-inertial device, where the inertia of a rotating mass provides the workload. Unlike the gravitational system, the iso-inertial method can provide a greater force in the eccentric phase of an exercise, known as eccentric overload. Owing to the force-generating system of iso-inertial systems, high workloads can be applied for both phases of a given action compared to gravitational systems, where eccentric overload can only be achieved with external assistance. Another benefit of the iso-inertial method is that, when the load increases or fatigue appears, the execution speed is reduced but the action is not interrupted (10). This allows a more fluid movement (i.e., self-adaptive resistance) than gravitational systems’ interrupted movements.

Maroto-Izquierdo et al. conducted a systematic review to evaluate the effects of iso-inertial training among athletes and healthy subjects, reporting an increased muscle strength, power and size (11). More recent studies have also focused on older adults. These studies showed improvements in postural control, maximal isometric strength, or isokinetic power in participants who used iso-inertial systems compared to those who trained with free weights (12,13). Iso-inertial training has also shown positive effects on metabolic variables such as lipid profiles or maximal volume oxygen consumption (14,15). However, no study has compared the effects of iso-inertial and gravitational training on power when performing functional exercises and on physical performance variables in middle-older adults.

The main objective of this study was to evaluate the effectiveness of an iso-inertial resistance training program on eccentric power compared to that of the same program executed with gravitational resistance in physically active middle-older adults. The secondary objective was to compare these two programs in concentric power, physical performance, and risk of falls.

## Methods

### Trial design and setting

This was a parallel-group, randomised controlled trial (allocation ratio 1:1) with a blinded outcome assessment performed at the Espai Esport Wellness Center gymnasium (Granollers, Spain).

We followed the Consolidated Standards of Reporting Trials (CONSORT) to produce this report (16).

### Participants

We recruited adults aged 57 years or older who were physically active. By “physically active” we considered that they were enrolled in the gymnasium mentioned above and used their facilities with varying frequency. We excluded participants with osteoarticular or acute musculoskeletal injuries or systemic neurodegenerative diseases. Recruitment was performed in collaboration with the administrative staff of the gymnasium, who contacted all members over 57 years old and invited them to participate.

### Interventions

Participants had to complete a 6-week program (2 sessions/week separated by at least 48 hours) using either an iso-inertial or a gravitational resistance device.

The sessions started with a warm-up that consisted of 1) 4 minutes of moderate aerobic exercise (treadmill, elliptical bike, or static bike), 2) active stretching exercises for 6 seconds each (hip adductors, hamstrings, gastrocnemius, quadriceps, and gluteus), and 3) 6-8 repetitions of each of the three intervention exercises without any resistance so that participants became familiar with them.

Then, regardless of the assigned group, participants had to perform the same three intervention exercises (forward lunge, side lunge, and forward lunge with row (see Figure 1). For the forward lunge and the forward lunge with row, the participant was placed frontally on the training device, whereas for the side lunge the participant was placed sideways on the device (homolaterally on the scrolling limb). For the forward and side lunge, we used a weight belt placed on the waist. For the forward lunge with row, we used a hand grip and the pull was performed with the upper limb homolateral to the displaced lower limb. Each exercise was performed with both limbs (Figure 1).

**Figure 1:**
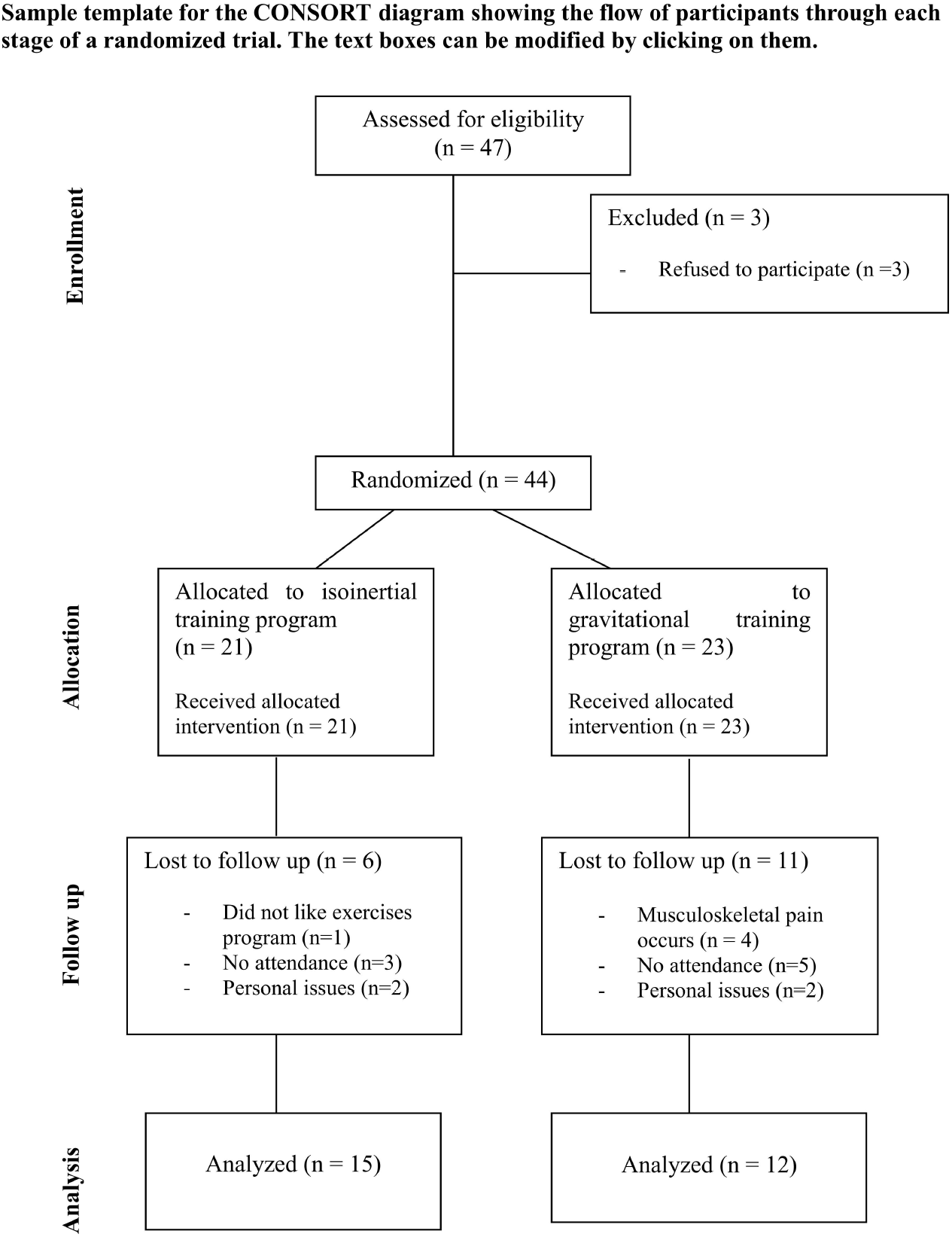
Intervention exercises: a) forward lunge, b) side lunge, and c) forward lunge with row.

After several repetitions to initiate the flywheel impulse, the exercises were performed to accelerate the rotation of the flywheel in the concentric action, and decelerate it in the eccentric action. The study investigators instructed participants to perform the exercises at a given intensity according to the Borg Rating of Perceived Exertion (RPE) scale (0-10, where 0 means no exertion at all and 10 means maximal exertion) (11). The volume, intensity, and difficulty increased over time (see Table 1). The load was identical for all participants, but each participant adjusted the execution speed of each exercise to achieve the desired RPE. At weeks 4 and 6, the exercises included variations in the directions and distances of the lunges that were marked on the floor.

**Table 1.** Progression of the intervention program.

| Sessions | Load | Intensity |
| --- | --- | --- |
| From 1 to 4 | 2 sets of 6 to 8 repetitions | RPE 3 to 5 |
| 5 and 6 | 3 sets of 10 repetitions | RPE 3 to 5 |
| 7 and 8 | 3 sets of 10 repetitions* | RPE 3 to 5 |
| 9 and 10 | 3 sets of 10 repetitions* | RPE 7 to 8 |
| 11 and 12 | 3 sets of 10 repetitions* | RPE 7 to 8 |

*RPE, rate of perceived exertion; \* lunges performed with different lengths and directions*

The iso-inertial group used the Nessinertial® conical pulley with 6 inertial loads. The rope regulator was placed at the lowest possible position so that the rope rolled in a large diameter, providing more speed but less drag. The participant was placed at a distance from the device determined individually when the traction string reached the maximum tension at the start of the execution of each exercise. The gravitational group used an MFT CSX-5000 device (weight: 3,75 kg). Participants were instructed not to perform any other resistance training for the lower limbs during the intervention period.

All participants were supervised by one of the study investigators or the center’s instructors until the participants were shown to be autonomous. From that moment, the sessions were performed without supervision. Additionally, participants were provided with documentary information through images and videos of the exercises. In case of doubts, they could contact the investigators by phone.

### Outcomes

Outcomes were assessed at the gymnasium’s facilities before the intervention (T0) and immediately after the completion of it (T1). The starting limb for each exercise was randomised for each subject.

The primary outcome was the power in the eccentric phase of each exercise. It was measured with both iso-inertial and gravitational resistance devices using a rotatory and a linear encoder (Chronojump Boscosystem®)(17), respectively. We used the free software Chronojump to record the data. This software is associated with the open hardware Chronopic V.3.

The secondary outcomes were 1) *power in the concentric phase* of each exercise, measured with both iso-inertial and gravitational devices; 2) *physical performance*, measured using the Short Physical Performance Battery (SPPB) (18), which consists of a *balance test*, a *speed of march test, and a five times sit-to-stand test*; and *3) risk of falls*, assessed through the *Get Up and Go test* (19). These tests are explained in Supplementary File 1.

We also collected anthropometric and sociodemographic variables: date of birth, weight (kg), height (cm), sex (man/woman), and work status (working/retired).

### Sample size

We performed the sample calculation using R statistical software (“pwr” package). Based on a previous study that compared iso-inertial and gravitational resistance training protocols in athletes using an iso-inertial device (10), we estimated an effect size and a standard deviation of 202 W and 153’97 W, respectively, for the primary outcome. No similar studies were found that used a gravitational system for power assessment. Assuming alpha = 0.05, a beta = 0.1, and a 10% dropout rate, the sample size was 30 participants (15/group).

### Randomisation (Sequence generation, Allocation concealment mechanism, Implementation) and blinding

We performed randomisation in blocks of 4 and stratified by sex and age (57-63, 64-70, 71 or more). The randomisation sequence was matched by the administrative staff of the center with the participants’ membership numbers according to the participants’ recruitment order. In this way, the participants and researchers were not aware of the allocation until the training program started. Given the nature of the intervention, neither participants nor investigators monitoring the training protocols may be blinded to the allocation. However, the outcome assessors did not know the assignment of each participant.

### Statistical methods

For the statistical analysis, we used R software. Initially, we presented descriptive data of all subjects who were randomised and who completed the study. To analyse the results for the study outcomes, we used linear regression models where the dependent variables were the change scores of each outcome, defined as the T1 minus the T0 value, and the independent variables were the allocation group (iso-inertial or gravitational), the limb, and some demographic variables (age and sex). We checked different model application assumptions (linearity, homoscedasticity, normality, and independence). As we observed that the characteristics of the lost to follow-up participants were similar to those of the participants who completed the study, we performed a complete case analysis. We reported the adjusted difference of means between groups along with its 95% confidence interval and the p-value of the comparison. The significance level was 0.05. The R scripts have been publicly shared (20).

Results

Between 10 November 2023 and 28 February 2024, we recruited 47 participants. Three of them refused to participate, therefore we randomised 44 participants: 21 to the iso-inertial (IN) and 23 to the gravitational (GR) training group. Seventeen patients were lost to follow-up (n=6 IN group, n=11 GR group). The main reasons for these were 1) failure to attend training sessions (n=8) and 2) occurrence of personal (n=4) or musculoskeletal problems (n=4). Finally, 27 participants completed the program (n=15 IN group, n=12 GR group) (Figure 2).

**Figure 2:**
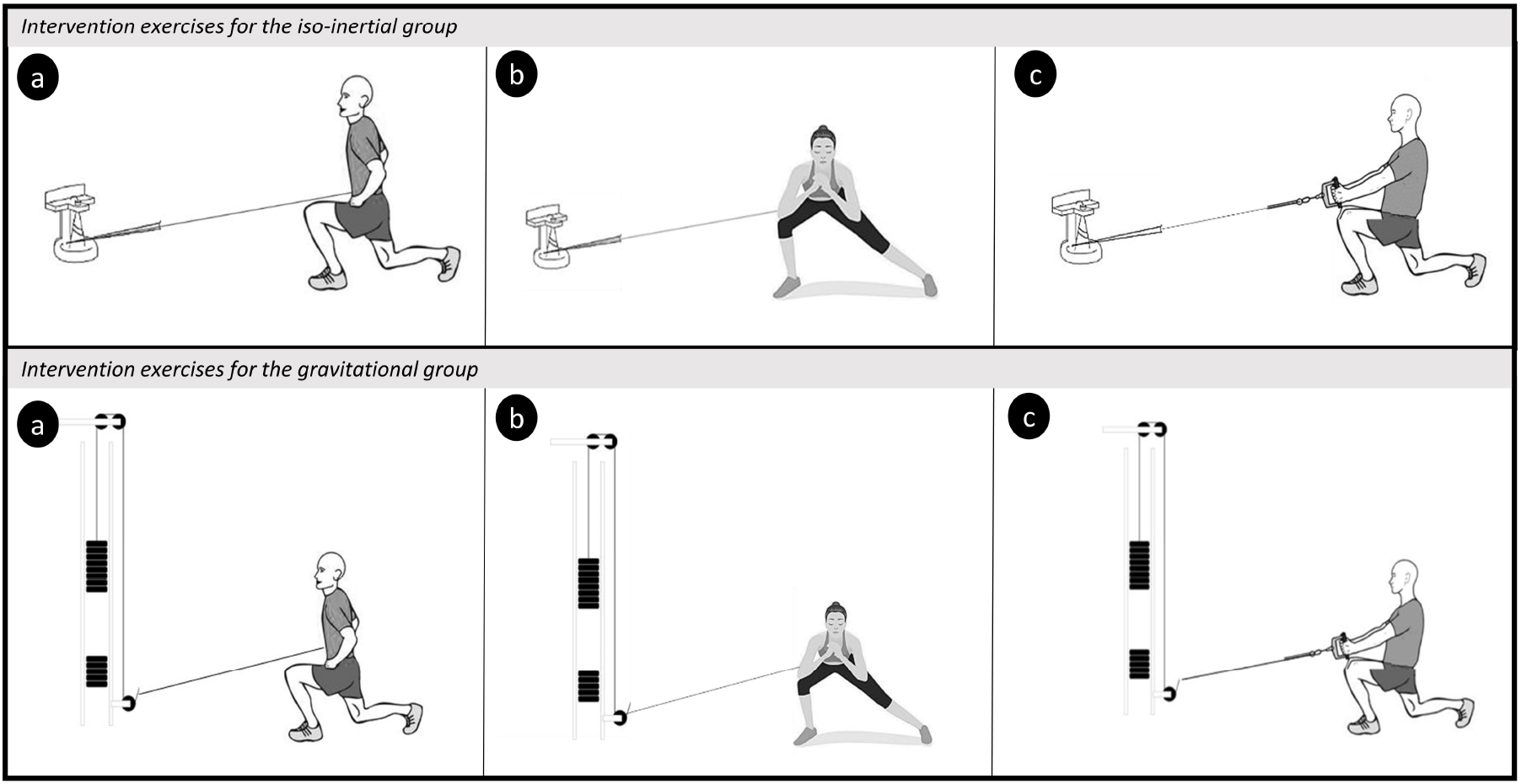
CONSORT flow diagram.

The mean (SD) age of participants was 63.81 (6.65) years, 56.81% (n=25) of the participants were females and 43.18% (n=19) were males. The baseline characteristics of all participants were similar to the characteristics of those who completed the study (Table 2).

**Table 2.** Baseline characteristics of participants.

|  |  | Total participants included (n=44) |  | Participants with full program completed (n=27) |  |
| --- | --- | --- | --- | --- | --- |
|  |  | IN group (n=21) | GR group (n=23) | IN group (n=15) | GR group (n=12) |
| **Age** | *Years (mean, SD)* | 65, 6,36 | 62,74, 6,87 | 64,53, 6,9 | 62,25, 6,61 |
| **Gender** | *Female (% ,n)* | 61,90%, 13 | 52,17%, 12 | 53,33%, 8 | 66,67%, 8 |
|  | *Male (% ,n)* | 38,1%, 8 | 47,83%, 11 | 46,67%, 7 | 33,33%, 4 |
| **Weight** | *Kilograms (mean, SD)* | 68,18, 10,28 | 73,09, 11,18 | 68,41, 9,35 | 70,67, 0,59 |
| **Height** | *Meters (mean, SD)* | 1,65, 0,08 | 1,69, 0,08 | 1,65, 0,06 | 1,67, 0,1 |
| **Status** | *Active (% ,n)* | 47,62%, 10 | 65,22%, 15 | 53,33%, 8 | 66,67%, 8 |
|  | *Retired (% ,n)* | 52,38%, 11 | 34,78%, 8 | 46,67%, 7 | 33,33%, 4 |

GR, gravitational; IN, iso-inertial; SD, Standard deviation

### Primary outcome

The improvement in the power in the eccentric phase evaluated with the iso-inertial system was greater in the IN group for all exercises (Table 3). However, there were statistically significant differences only for the *side lunge*. For the *forward lunge*, the mean power in the eccentric phase at T1 was 42.67 W in the IN group and 35.59 W in the GR group. The difference between groups was 3.99 W (95% CI: -3.99 to 11.33, p=0.28). For the *side lunge*, the mean eccentric power was 51.06 W and 40.22 W in the IN and GR groups, respectively (between-group difference: 8.50 W, 95% CI: 2.13 to 14.87; p=0.01). The mean eccentric power for the forward lunge with row was 128.39 W and 102.41 in the IN and GR groups, respectively (between-group difference: 14.07 W, 95% CI: -2.07 -to 30.20, p=0.09).

**Table 3.** Intra and inter-group analysis of power changes in the eccentric phase.

| Power in eccentric phase (W) |  | GR Group |  | IN Group |  | Differences between groups | Significance |
| --- | --- | --- | --- | --- | --- | --- | --- |
| Evaluation system | Exercise | mean (SD) | mean (SD) | mean (SD) | mean (SD) | dif_adjusted (95% CI) | p-value |
|  |  | T0 | T1 | T0 | T1 |  |  |
| Iso-inertial | Forward lunge | 19,12 (9,37) | 35,59 (21,11) | 21,14 (9,52) | 42,67 (15,78) | 3,99 (-3,35 to 11,33)* | 0,28 |
|  | Side lunge | 25,52 (12,21) | 40,22 (15,25) | 26,75 (12,04) | 51,06 (19,37) | 8,5 (2,13 to 14,87)* | 0,01 |
|  | Forward lunge with row | 65,89 (45,58) | 102,41 (62,81) | 71,89 (40,08) | 128,39 (51,49) | 14,07 (-2,07 to 30,2)* | 0,09 |
| Gravitational | Forward lunge | 21,52 (6,36) | 26,08 (8,03) | 21,53 (6,50) | 24,99 (5,41) | -0,34 (-2,86 to 2,18) | 0,79 |
|  | Side lunge | 19,11 (4,26) | 21,75 (4,22) | 19,39 (4,07) | 23,03 (4,40) | 0,56 (-1,53 to 2,65)* | 0,59 |
|  | Forward lunge with row | 42,37 (16,38) | 60,08 (27,24) | 42,41 (18,57) | 55,58 (16,79) | -2,99 (-13,48 to 7,51) | 0,57 |

W, watts; GR, gravitational; IN, iso-inertial; SD, standard deviation; CI, confidence interval; \*favors iso-inertial group

Both groups had similar power changes in the eccentric phase evaluated with the gravitational system (Table 3). For the *forward lunge*, the mean eccentric power was 24.99 W and 26.08 W in the IN and GR groups, respectively (between-group difference: -0.34 W, 95% CI: -2.86 to 2.18, p=0.79). For the *side lunge*, the mean eccentric power was 23.03 W in the IN and 21.75 W in the GR group (between-group difference: 0.56 W, 95% CI: -1.53 to 2.65, p=0.59). The *forward lunge with row* showed a mean eccentric power of 55.58 W and 60.08 W in the IN and GR groups, respectively (between-group difference: -2.99 W, 95% CI: -13.48 to 7.51, p=0.57).

### Secondary outcomes

The changes in concentric muscle power evaluated with both iso-inertial and gravitational systems were similar in both groups (Table 4). There were also no remarkable between-group differences in physical performance and risk of falls (Table 5).

**Table 4.** Intra and inter-grup analysis of power changes in the concentric phase.

| Power in concentric phase (W) |  | GR Group |  | IN Group |  | Differences between groups | Significance |
| --- | --- | --- | --- | --- | --- | --- | --- |
| Evaluation system | Exercise | mean (SD) | mean (SD) | mean (SD) | mean (SD) | dif_adjusted (95% CI) | p-value |
|  |  | T0 | T1 | T0 | T1 |  |  |
| Iso-inertial | Forward lunge | 21,74 (9,18) | 39,51 (24,24) | 26,55 (10,74) | 42,29 (14,45) | -3,23 (-10,65 to 4,19) | 0,39 |
|  | Side lunge | 25,49 (12,40) | 41,09 (19,00) | 28,65 (12,66) | 48,02 (17,43) | 1,70 (-6,08 to 9,48)* | 0,66 |
|  | Forward lunge with row | 72,19 (48,28) | 106,71 (60,33) | 79,91 (44,43) | 126,10 (50,11) | 5,65 (-10,12 to 21,43)* | 0,47 |
| Gravitational | Forward lunge | 19,14 (5,43) | 21,74 (4,74) | 19,91 (5,27) | 21,92 (5,09) | -0,32 (-2,70 to 2,06) | 0,79 |
|  | Side lunge | 17,43 (3,61) | 18,42 (3,46) | 17,45 (3,04) | 19,60 (2,81) | 0,97 (-0,25 to 2,18)* | 0,12 |
|  | Forward lunge with row | 43,44 (20,31) | 71,44 (48,66) | 52,60 (27,81) | 82,04 (37,79) | 3,26 (-16,72 to 23,24) | 0,74 |

W, watts; GR, gravitational; IN, iso-inertial; SD, standard deviation; CI, confidence interval; \*favors iso-inertial group

**Table 5.** Intra and inter-grup analysis of physical fitness and risk of falls changes.

| Physical fitness |  | GR Group |  | In Group |  | Differences between groups | Significance |
| --- | --- | --- | --- | --- | --- | --- | --- |
| Test | Outcome | mean (SD) | mean (SD) | mean (SD) | mean (SD) | dif_adjusted (95% CI) | p-value |
|  |  | T0 | T1 | T0 | T1 |  |  |
| SPPB | Score (0 to 12) | 10,92 (1,16) | 11,67 (0,65) | 10,85 (0,80) | 10,92 (1,32) | 0,66 (-0,38 to 1,69) | 0,20 |
|  | Balance (sec) | 3,83 (0,58) | 4,00 (0,00) | 4,00 (0,00) | 3,92 (0,28) | 0,25 (-0,13 to 0,63) | 0,19 |
|  | Walking speed (sec) | 3,18 (0,59) | 3,09 (0,46) | 3,18 (0,62) | 2,86 (0,46) | 0,22 (-0,19 to 0,63)* | 0,28 |
|  | FTSST (sec) | 12,25 (2,01) | 10,84 (2,18) | 12,66 (2,08) | 11,30 (2,36) | -0,24 (-1,42 to 0,95) | 0,68 |
| Risk of falls |  | GR_Group |  | In_Group |  | Differences between groups | Significance |
| Test | Outcome | mean (SD) | mean (SD) | mean (SD) | mean (SD) | dif_adjusted (95% IC) | p-value |

|  |  | T0 | T1 | T0 | T1 |  |  |
| --- | --- | --- | --- | --- | --- | --- | --- |
| GUG | Seconds | 8,29 (1,60) | 8,04 (1,02) | 8,33 (1,28) | 7,67 (1,20) | 0,37 (-0,77 to 1,52)* | 0,51 |

GR, gravitational; IN, iso-inertial; SD, standar desviation; CI, confidence interval; SPPB, Short Physical Performance Battery; sec, seconds; FTSSST, five times sit to stand test; GUG, Get up and go test; \*favors to iso-inertial group.

## Discussion

Our results indicated that iso-inertial training led to greater power gains in the eccentric phase than gravitational training when power was assessed with the iso-inertial system. However, with the gravitational system evaluation, the two types of training performed similarly. In addition, we observed no differences in the concentric power, physical performance, and risk of falls. This is the first study in middle-older adults to 1) compare the effectiveness of iso-inertial and gravitational resistance training on the power of a coordinative action, 2) separately analyse the power in the eccentric and concentric phases of each exercise, and 3) assess power using both an iso-inertial and a gravitational device regardless of the participant’s training group.

Despite the importance of muscular power in middle-older adults (4), only two studies have compared iso-inertial and gravitational training using this parameter. Floreani et al. evaluated the maximal explosive power when performing a *leg press* (force platform), observing that both training approaches led to similar gains (+11.1% IN, +13.4% GR)(7). In a similar study, Sañudo et al. reported a 63% increase in power output in the IN group (with no data from the GR group) (21). These findings are difficult to compare with ours for three reasons: 1) different evaluation systems, 2) different training protocols, and 3) no concentric-eccentric phase differentiation when measuring power. First, the improvements by Floreani et al. might be similar to ours when power was assessed using the gravitational system, but very different from those from the iso-inertial system assessment, where our gains in both groups were much greater and favoured the IN group. These results are consistent with the fact that they evaluated power with a noniso-inertial system. However, Sañudo et al. also evaluated power using a noniso-inertial system and their results are remarkably different from ours. In general, we believe that the iso-inertial system evaluations reflect the power gains more accurately than the gravitational evaluations because of the eccentric overload generated by the device. Therefore, we suggest using iso-inertial devices to evaluate the changes generated by eccentric overload training. Second, the two studies above proposed a training program based on a single analytical exercise (squat or leg extension). Our protocol is functional because it involves trunk stabilization and the main upper and lower extremity muscle groups, and combines resistance and high-velocity exercises. This protocol aligns with current position statements and consensus guidelines for physical activity in older adults, which recommend a multimodal prescription that includes aerobic, strength balance, and flexibility (22). In this regard, training protocols should always include functional actions with maximal transference to middle-older adults’ activities of daily living. Third, the two studies above evaluate power only in absolute terms, making no difference between eccentric and concentric phases. Given the implications of eccentric actions for activities of daily living, a separate analysis of power in the eccentric phase should always be included in future studies that evaluate training programs in middle-older adults (23–25).

The IN group obtained greater power in the eccentric phase values than the gravitational group when power was assessed with the iso-inertial system. These results are consistent with the theoretical basis of iso-inertial training, which allows maximal concentric and eccentric muscle actions, with brief episodes of eccentric overload (10). Previous studies have reported that the peak force generated in the eccentric phase of movement may be 15-30% greater than that produced in the preceding concentric action due to the elastic energy storage characteristics of the iso-inertial system (26,27). In this regard, animal studies have demonstrated that increased protein synthesis (vinculin, titin, and nebulin) is linked to physiological tissue adaptations and elastic properties in response to eccentric work (28).

No differences between groups were observed in eccentric power for the gravitational system evaluation. This might be because of the limited power generation during the eccentric phase when gravitational training is performed. Previous studies in athletes have reported improvements in power favoring iso-inertial training when comparing iso-inertial and gravitational training, however, both groups were evaluated only with the iso-inertial system (11,29–31). Using a single tool could attribute the results to the participants’ familiarity (and lack thereof) with the iso-inertial system. Assessing our participants with both systems allowed us to avoid this bias. Indeed, if the differences favoring the iso-inertial training group were a consequence of familiarisation with the training method, the GR group would have obtained more power on the gravitational system evaluation. However, the gravitational system evaluation showed almost no differences between the groups. This finding confirms that the eccentric overload generated by iso-inertial training generates true higher power values in the eccentric phase of the action.

Although there were no significant differences between groups, we found that all participants improved their physical performance and reduced their risk of falls. These results are in line with the systematic review of training programs in older adults that reported improvements in functional performance for all participants, with a small advantage for eccentric-based compared to concentric-based exercises (32). Another review reported improvements in unipedal balance in older adults who used an iso-inertial device compared to those who used weights (11). The authors attributed this response to increased tendon stiffness and neuromuscular transfer to the plantar flexors. Notably, these reviews are focused on analytical exercises. However, our protocol proposed a more functional approach. First, we used lunges because they involve complex functions such as deceleration of a limb, force absorption, and controlling movement against an external force (33). Notably, these functions are key components of an eccentric action (25). In addition, all exercises accentuated trunk stabilization, which should always be a factor to consider when developing interventions for middle-older adults. Some studies have reported that core stability training for older adults can also improve balance and coordination, and decrease the risk of falls (34).

On the other hand, few studies have compared the effects of performing the same functional exercises with different types of resistance. Madruga-Parera et al. evaluated the effectiveness of functional exercises for handball players that were biomechanically identical for the two study groups and only the type of resistance changed (35). Our protocol was based on this approach. We suggest that future studies incorporate this methodology when evaluating the differences between training methods to avoid performance biases in their results.

It is worth mentioning that the scales used for evaluating physical performance and risk of falls lacked sensitivity for the type of participants included, as we encountered a ceiling effect. For example, the participants’ mean baseline SPPB score was 10.88 out of 12 points, indicating a great physical condition. A similar ceiling effect was noted in another study, that claimed that the Berg scale could not predict the risk of falling at high levels of balance ability (36). As other studies do, we suggest that tests and scales originally built for middle-older adults should be modified for physically active middle-older adults.

### Strengths and limitations

The strengths of this study include 1) the randomised controlled trial design, 2) the pragmatic design, which is ideal for testing the effectiveness of interventions under real-life conditions, 3) the outcome evaluations with two different systems to avoid familiarisation effects, 4) the functional training program, and 5) the inclusion of identical exercises for the two groups.

This research also has some limitations. First, 38% of participants were lost to follow-up. However, their characteristics were similar to those of those who completed the study. Future research should place more emphasis on closer monitoring of interventions. Additionally, we did not collect data on participant satisfaction, which would have allowed us to determine whether the greater losses in the GR group were due to dissatisfaction with the training device. Finally, we did not measure further physiological parameters involved in functional and structural adaptations. Future studies could include these parameters to explore the underlying mechanisms behind the effects of iso-inertial and gravitational training.

### Clinical implications

This study helps improve the understanding of the effects of iso-inertial and gravitational resistance training, leading clinicians to recommend more effective training programs for older adults. Additionally, we provide strong empirical data that support orienting resistance training towards complex exercises involving complex coordinative actions (rather than analytical exercises) with a transfer to daily activities such as walking or going up and down stairs. This approach may increase older adults’ autonomy and therefore promote healthier longevity. Finally, we warn about the ceiling effect of the current clinical evaluation tests and suggest avoiding their use in the physically active middle-older adult population. Instead, we propose to use power assessments in clinical practice to monitor the improvements associated with resistance training programs.

### Conclusions

A 6-week iso-inertial training program led to greater power in the eccentric phase gains than gravitational training when power was assessed with an iso-inertial system. However, there were no differences between the two methods for the gravitational system evaluation or for concentric power, physical performance, and risk of falls. Regardless of the training system, the resistance training program remarkably improved the results for all outcomes.

## Other information

## Acknowledgements

The authors warmly thank the staff of Espai Esport Wellness (Granollers, Spain) for allowing us to carry out this project in their facilities and helping us recruit participants and monitor the intervention.

## Contributors

Conceptualization: AC, DR, MMP, DB. Writing – original draft: AC, DB. Writing – review & editing: AC, DB, DR, MMP, VZ and FD. Methodology, Software, Data Curation and Formal Analysis: DB and VZ. Resources: DR, MP. Investigation: AC, DB, DR, FD. Supervision and project administration: AC, DB.

## Patient and public involvement

Patients and/or the public were not involved in the design, conduct, reporting, or dissemination plans of this research.

## Ethics approval

This study was approved by the Drug Research Ethics Committee of the Universitat Internacional de Catalunya (Code: FIS-2023-03). The protocol can be found in Supplementary File 2. Participants provided informed consent to participate in the study before taking part.

## Provenance and peer review

Not commissioned; externally peer-reviewed.

## Data availability statement

Data were stored and encoded in three Excel files to protect confidentiality before, during, and after the study. The first of these files contains the personal data of the participants (name and surnames, age, and sex), is accessible only to the administrative staff of the center, and served only to manage the recruitment of the participants. The second and third files are managed entirely by the research team and contain the registry of all the study outcomes. In these files, participants were identified by a code. Only the research team can relate the data collected in the study to the identity of the participants, which will not be available to anyone except in the event of a medical emergency or legal requirement. These two files have been shared in a public repository (20).

## Study Registration

This study was registered at Clinicaltrials.gov (NCT06160089).

## Work address

C/ de Josep Trueta, 08195 Sant Cugat del Vallès, Barcelona, Spain.

## Supplementary Files

**Supplementary File 1:** Explanation of the Short Physical Performance Battery and Get Up and Go tests.

**Supplementary File 2:** Study protocol approved by the Ethics Committee.

