## Supplementary File 1 for "Effectiveness of iso-inertial resistance training on eccentric and concentric power, physical performance, and risk of falls in physically active middle-older adults: a randomised controlled trial"

**Short Physical Performance Battery (SPPB)**

*Balance test.* It consists of standing in three different positions for 10 seconds (feet together, semi-tandem, and tandem with open eyes). For each maintained position, 1 point is given, except for the tandem, where the maximum score is 2. Therefore, the maximum score for this test is 4 points.

*Speed of march test.* It measures the time the subject takes to travel 4 metres indicated by two marks on the floor. Timekeeping was performed during this distance, but a space of one-meter acceleration before the start point and one-meter deceleration after the endpoint was allowed. The highest mark of two attempts was recorded. One point is administered if the attempt takes more than 8.70 seconds, 2 points if it takes 6.21 to 8.70 seconds, 3 points if it takes from 4.82 to 6.20 seconds, and 4 points if it takes less than 4.82 seconds.

*Five times sit to stand test.* It focuses on the lower limbs functionality assessment*.* The participant is asked to perform 5 times the following action: rising from a chair at the maximum possible speed without the help of the lower limbs and sitting down again. The time taken to complete one single attempt of this action is recorded. 0 points are awarded if the participant is unable to perform 5 repetitions or if the attempt takes more than 60 seconds, one point if it achieves it with a time of between 16.7 and 60 seconds, 2 points from 13.7 to 16.69 seconds, 3 points from 11.2 to 13.69 seconds and 4 points when the time is equal or less than 11.19 seconds. Combining the three tests from this battery, the participant can obtain a maximum score of 12 points. If the total score is below 8 points, the participant is considered to have a low physical aptitude.

***Get up and go* (GUG) test**

It is commonly used test to assess the basic mobility of older adults and possible gait and balance disorders associated with the risk of falls. This test starts with the participant sitting on a chair. When the timing begins, he or she has to walk 3 meters at a normal speed, turn around a cone and walk back at the same speed to sit back on the chair. The score obtained in this test corresponds to the time (in seconds) taken to complete the whole action. The risk of falls is considered normal if the attempt takes less than 10 seconds. If it takes between 11 and 13, the participant is considered to have a slight disability in mobility, and if it takes more than 13 seconds, the risk of falls is considered high.
