## Supplementary File 2 for "Effectiveness of iso-inertial resistance training on eccentric and concentric power, physical performance, and risk of falls in physically active middle-older adults: a randomised controlled trial"

**Effectiveness of iso-inertial resistance training on the muscle power of the lower limbs, physical aptitude, and risk of falls in physically active older adults: a randomized controlled trial.** Cadellans-Arróniz, Aïda^1^; Romero-Rodríguez^1^, Daniel; Madruga-Parera^1^, Marc; Ortega-Cebrian, Silvia, Marc; Dantony Flora, Blanco de Tena Dávila, David^1^

^1^Universitat Internacional de Catalunya, Department of Physiotherapy, Sant Cugat del Vallès, Barcelona, Spain

**Abstract**

**Background:** Strength training is effective for combating sarcopenia, risk of falls and fragility in the elderly. The gravitational strength method is the most traditionally used approach in community health. However, the workload applied is limited to concentric muscle capacity. In contrast, the iso-inercial method causes an overload of the eccentric phase that has been proven effective in older adults. However, no study has compared the effects of eccentric and concentric training on muscle power and other functional variables in older adults.
**Objectives**: To evaluate the effectiveness of an iso-inertial resistance program on eccentric and concentric muscle power of the lower limbs, physical fitness and risk of falls compared to the same program executed with gravitational resistance in physically active older adults.
**Methods:** Randomized controlled trial conducted at the gymnasium Espai Esport Wellness Center (Granollers, Catalonia). The sample will be 30 physically active adults (age ≥60), excluding participants with acute musculoskeletal injuries, systemic or neurodegenerative diseases. Intervention will be a 6-week program (2 sessions/week) using either an iso-inertial or a gravitational device. We will build generalised linear models to evaluate the effects of interventions on muscle power during eccentric phase (primary outcome, measured with Chronojump BoscosystemR), muscle power during concentric phase, physical aptitude, and risk of falls (secondary outcomes). All outcomes will be assessed before and after the intervention. Participants will have to sign the informed consent form.
**Discussion:** This study will explore whether a new approach of strength training is more effective for a population where preserving functional capacities is essential for longevity. Bringing older adults into an active environment could reduce the risk of physical and mental diseases, decreasing public health costs. Also, the results will generate knowledge that will guide physiotherapy care practice, promote coordination between health professionals and open future research lines focused on eccentric work.

**Key words:** resistance training, muscle strength, muscle power, iso-inertial training, lower extremity, exercise, accidental falls, aged.

**Background:** Actions aimed at promoting healthy ageing in the older adults are key to slow down the physiological progressive loss in skeletal muscle mass, quality and function as the person ages. These regressions affect the individual's ability to carry out daily life activities. In this respect, muscle strength and power are reduced during aging, which has a negative impact on the functional capacity and quality of life of older adults. In fact, recent research has identified a significant decrease in muscle strength of 1-1.5% each year from the age of 50 (1)(2). On the one hand, muscle strength is defined as the ability to generate intramuscular tension when facing a resistance, regardless of whether it generates movement. On the other hand, muscle power is defined the maximum amount of force one can generate during a specific movement at a specified velocity (3). Muscle power is considered a predictor of functional capacity, as it is associated with everyday activities such as climbing stairs, standing up from a chair or walking (4). This fact has justified the use of strength and power training in different studies to prevent the risk of falls, improve the balance or the walking capacity in older adults (5). Previous studies report that muscle power is diminished to a greater extent than strength over time (2).

Resistance training (RT) is considered one of the main strategies to prevent the functional capacity decrease. This type of training produces adaptations at different structural levels (6) and has demonstrated its effectiveness in combating age-induced muscle atrophy (sarcopenia), risk of falls and fragility. It has also been shown to improve cardiovascular health (3). Previous studies have noted statistically significant differences in favor of eccentric training, when comparing eccentric and concentric RT. Specifically, higher peak strength with lower muscle activation and lower metabolic cost, increased muscle mass and higher jumping performance have been observed (7). Thus, the values of muscle strength and power resulting from an eccentric RT are physiologically superior to those that can be obtained when performing concentric RT (1)(7).

The most traditionally used RT method in the field of community health is the one known as gravitational, in which a resistance is opposed through free weights or by blocks or disks (cable machines). One of the main restrictions of this method is that the workload applied during the shortening-longing cycle of a repetition is limited to concentric muscle capacity and does not allow a progression of the eccentric workload. This fact limits the potential of this method for generating the improvements associated with eccentric training mentioned above (1). In contrast, the iso-inertial (ISI) training method is based on the application of resistance generated by an iso-inertial device, where the workload is provided by the inertia of a rotating mass. Unlike the gravitational system, the ISI method can provide a resistance workload in the eccentric phase that is proportional to the concentric phase. Thanks to that, a high workload can be applied for both concentric and eccentric cycles. In addition, it allows to perform the established repetitions without having to stop the scheduled training program. This is due to the fact that, when the load increases or the fatigue appears, only the execution speed (7) is reduced.

Different studies indicate that eccentric overload training protocols (i.e., a higher force in the eccentric phase than the concentric phase of a repetition) cause higher muscle-level improvements compared to other actions. Because of its force-generating system, the iso-inertial mechanism is ideal for eccentric overload training. compared to gravitational systems where eccentric overloading can only be performed with external assistance. In addition, the ISI system allows a fluid movement compared to the traditional system where the movement is executed in an interrupted way.

Exposure to prolonged eccentric work has been studied in sporting contexts showing an increase in muscle power in different ranges of motion and a decrease in muscle injuries. Maroto-Izquierdo et al. conducted a systematic review and meta-analysis to assess the effects of iso-inertial work with eccentric overload on sports population (8). Their results indicate improvements in muscle hypertrophy, neuromuscular functions, increased power and decreased sprint time. The authors attribute these improvements not only to eccentric overload work, but to the application of an accommodated resistance that provides an optimal workload for the user, thanks to the ISI method. Although ISI training has been extensively studied in young athletic populations, recent studies have been published that evaluate its effect in older adults. These studies show promising results, as ISI would improve postural control (5) and the maximum isometric force (4). Other studies have shown improvements in metabolic variables such as lipid profile or maximum oxygen volume consumption (2). However, no study has been found that compares the effects of these two types of training on muscle power and other relevant functional variables in older adults.

**Justification:** With the progressive increase in life expectancy of the population, attention to ageing has gained special interest in recent years. Physiological ageing causes changes in physical and cognitive functions that have an impact on the quality of life of older people. More concretely, some of the physical changes that have a greater impact on the activities of daily living are the loss of muscle strength and power. Studies have shown that physical exercise helps mitigate these physiological changes. More concretely, the benefits of eccentric work on neuromuscular adaptations in strength resistance training have been extensively described in younger populations. Eccentric work is especially relevant as it is directly related to actions such as walking downstairs or getting up from a chair. However, the effects of eccentric strength work on muscle power in older adults have not been deeply explored. It is also unknown how the potential neuromuscular improvements of older adults could have a transfer on their functional abilities. This is of particular importance as the improvement in their functional abilities would have a direct impact on their autonomy and quality of life. Furthermore, the development of the proposed project in a gymnasium is relevant from a socio-community point of view. Doing physical exercise in that context helps the elderly improve their social integration, which avoids isolation and improves self-esteem.

**Hypothesis:**

The main hypothesis is that iso-inertial resistance training program is more effective in improving the eccentric muscle power of the lower limbs compared to the same program executed with gravitational resistance in physically active older adults.

The secondary hypotheses are that 1) the isoinertial resistance training programme is more effective in improving concentric lower limb muscle power, physical fitness (balance, gait speed and lower limb functionality), and risk of falls compared to the same programme performed with gravitational resistance in physically active older adults, and 2) there are differences between men and women in the results of the variables analysed.

**Objectives:**

The main objective of this study is to evaluate the effectiveness of an iso-inertial resistance training program on the eccentric muscle power of the lower limbs compared to the same program executed with gravitational resistance in physically active older adults.

The secondary objectives are 1) to evaluate the effectiveness of an iso-inertial resistance training program on the concentric muscle power, physical fitness (balance, walking speed and functionality of the lower limbs), and risk of falls compared to the same program executed with gravitational resistance in physically active older adults, and 2) to assess the differences between men and women for each of the previous objectives.

**Methods:**

**Trial design:** This is a parallel group, randomized controlled trial (allocation ratio 1:1) with blinded outcome assessment where participants will be randomly assigned to one of the two study groups (iso-inertial or gravitational resistance training).

**Population:** Our sample will consist of adults aged 57 or more who are physically active, users of Espai Esport Wellness Center gymnasium where the study will be carried out (Passeig Conca del Besòs, 12, 08403 Granollers, Vallès Oriental, Catalonia, Spain). By “physically active”, we consider that they are enrolled in the gymnasium mentioned above and that they make use of their facilities with varying frequency in the months prior to the start of the study. We will exclude participants with osteoarticular or acute musculoskeletal injuries, systemic diseases (hepatic, pulmonary or cardiac renal failure, or chronic infectious diseases) or neurodegenerative diseases. The recruitment of participants will be done in collaboration with the administrative staff of the Espai Esport Wellness Center, which will contact all the members of the center who are over 60 years old and will offer them to participate in the study.

**Interventions:** Participants will have to complete a 6-week program with 2 sessions per week (separated by at least 48 hours) using either an iso-inertial or a gravitational device.

*Warm-up.* Before each training session, participants will perform a warm-up that consists of 1) moderate aerobic exercise (treadmill, elliptical bike, or static bike) for 4 minutes, 2) active stretching exercises with eccentric tension for 6 seconds (hip adductors, ischiosurals, gastrochronemis, quadriceps, and gluteus), and 3) one series of 6-8 repetitions of the three main exercises (see Annex 1) without any resistance (9). This part of the protocol will take place in the fitness room of the gymnasium.

*Intervention.* Regardless of the assigned group, participants will have to perform three exercises (front lunge, side lunge, and front lunge with hand grip pull that will be the basis of each session. For the front lunge and the front lunge with hand grip pull, the participant will be placed frontally on the training device (inertial or gravitational). Whereas for the side lunge the participant will be placed sideways on the device (homolateral on the scrolling limb). For front and side lunge, a weight shift belt will be used, which will be placed on the waist. For the frontal lunge with hand grip pull, the grip is performed with the upper limb homolateral to the displaced lower limb (see Annex 2). In addition, the volume and difficulty of execution of the exercises will increase over time (Annex 1). The study investigators will instruct the participants to perform the exercises at a given intensity according to the Borg Rating of Perceived Exertion (RPE) scale (10). The intensity will increase throughout the training program. Borg’s scale allows to measure the subjective perception of physical activity intensity and goes from 0 to 10, where 0 means "no exertion at all" and 10 means "maximal exertion". In the first 4 weeks, the exercises will have to be performed at a 3-5 RPE (moderate-hard), while a RPE of 6-8 (very hard) will be requested in the last two weeks. Starting from an identical load for all participants, the speed of execution of each exercise will be adjusted for each person to achieve the desired RPE. Each exercise will be performed with both limbs and the starting limb will be randomized for each subject.

Regardless of the allocation group, participants will receive the intervention individually and will be supervised by study investigators, who will verbally guide the participant and ensure the correct execution of each exercise. Each session will be performed in a separate room within the gymnasium where the training devices will be installed for the exclusive use of participants.

*Iso-inertial group.* The iso-inertial program will be executed using a Nessinercial® conical pulley device with 6 inertial loads. The participant will be placed at a distance from the device that will be determined individually when the traction string reaches the maximum tension at the start of execution of each exercise. In the conical pulley, the rope regulator will be placed at the lowest possible position. In this way, the rope will roll in a larger diameter, providing greater speed but less drag.

*Gravitational group*. The gravitational program will consist of the same set of exercises described above, but these will be executed with a traditional gravitational resistance system (cable resistance machine Titanium Strength THL Double Adjustable Pulley).

The training program will be interrupted if the participants get injured or declare that they would like to leave the study. Participants will not be able to perform any other type of resistance training for the lower limbs during the 6-week training period. However, they will be allowed to perform their usual aerobic exercise and resistance training for other muscle groups.

**Outcomes**

**Dependent variables**

Primary outcome: Muscle power during the eccentric phase measured with both iso-inertial and gravitational devices.

Secondary outcomes: 1) Muscle power during the concentric phase measured with both iso-inertial and gravitational devices. 2) Physical aptitude: Balance, walking speed, and lower limb functionality. 3) Risk of falls

**Independent variables:** Anthropometric and sociodemographic variables: date of birth, weight (kg), height (cm), sex (man/woman), work status (working/unemployed).

**Data collection methods and instruments.** Study outcomes will be assessed by two blinded evaluators in two moments: before the start of the intervention (T0) and immediately after the completion of the intervention (T1). Assessments will be conducted in the same separate room within the gymnasium where the training and evaluation material will be located. The instruments used to describe each of the outcomes are listed in Table 1 and described in detail below.

| **Variable (units)** | **Instrument** |
| --- | --- |
| Eccentric muscle power assessed with iso-inertial device (Watts) | Rotatory encoder in Proinertial Pulley Pro C3 (Chronojump Boscosystem®) |
| Concentric Muscle power assessed with iso-inertial device (Watts) | Rotatory encoder in Proinertial Pulley Pro C3 (Chronojump Boscosystem®) |
| Eccentric muscle power assessed with gravitational device (Watts) | Linear encoder (Chronojump Boscosystem®) |
| Concentric muscle power assessed with gravitational device (Watts) | Linear encoder (Chronojump Boscosystem®) |
| Balance (Seconds) | Tandem and semitandem** |
| Walking speed (Seconds) | 4 meters walking test** |
| Lower limb functionality (Seconds) | 5 times sit to stand test** |
| Risk of falls (Seconds) | Get up and go test |

*Table 1. Instruments and variables. The muscle power will be calculated as the average peak power of the best three consecutive repetitions in a series. **Tests contained within the Short Physical Performance Battery*

To evaluate the power during the eccentric and concentric phase of exercises, the encoder Chronojump BoscosystemR will be used. It is a validated (11) system used to evaluate kinematic data in actions associated with speeds. It is based on a chronograph that detects changes in the electric potential. The free Chronojump software associated with the open Chronopic V.3 hardware will be used and the log will be adjusted to the training mode being applied. In order to avoid a potential bias in the final assessments induced by the type of training assigned to the participant, participants will be assessed using both systems (iso-inertial and gravitational devices) before and after the intervention.

Short Physical Performance Battery will be used to evaluate physical fitness. More concretely, it assesses the physical ability of the lower limbs in older adults. This battery of tests has been validated (12) and has a good reliability. Its results are also considered indicators of frailty and disability. This battery is composed of the following three tests: **1) Balance** **test**: consists of holding three different positions in bipedal for 10 seconds (foots together, semi-tandem and tandem with open eyes). For each maintained position, 1 point is given, except for the tandem, where the maximum score is 2. Therefore, the maximum score for this test is 4 points. **2) Speed of march test.** It consists of measuring the length of time the subject takes to travel 4 metres marked by two marks on the floor. Timekeeping will be performed during this distance, but a space of one meter acceleration before the start point and one meter deceleration after the end point will be allowed. Two attempts are made and the highest mark is recorded. One point is administered if the attempt takes more than 8.70 seconds, 2 points if it takes from 6.21 to 8.70 seconds, 3 points if it takes from 4.82 to 6.20 seconds and 4 points if it takes less than 4.82 seconds. **3) Test of functionality of the lower limbs**. The participant is asked to perform 5 times the following action: rising from a chair at the maximum speed possible without the help of the lower limbs and sitting down again. One attempt is made and the time to complete it is measured. 0 points are awarded if the participant is unable to perform 5 repetitions or if the attempt takes more than 60 seconds, one point if it achieves it with a time of between 16.7 and 60 seconds, 2 points from 13.7 to 16.69 seconds, 3 points from 11.2 to 13.69 seconds and 4 points when the time is equal or less than 11.19 seconds. Combining the three tests, the participant can obtain a maximum score of 12 points. If the total score is below 8 points, the participant is considered to have a low physical aptitude.

The risk of falls will be assessed through the **Get up and go test.** This test is commonly used to assess the basic mobility of older adults and possible gait and balance disorders REF. These variables are particularly associated with the risk of falls (13). To start the Get up and go test, the participant will sit in a chair. When the timing begins, he or she will have to walk at a normal speed a distance of 3 meters, turn around a cone and walk back at the same speed to sit back in the chair. The score obtained in this test corresponds to the time (in seconds) it takes to complete the whole action. The risk of falls is considered normal if the attempt takes less than 10 seconds. If it takes between 11 and 13, the participant is considered to have a slight disability in mobility, and if it takes more than 13 seconds, the risk of falls will be considered as high. Independent variables will be recorded using a form designed for this purpose.

**Sample size:** We have performed the sample calculation using the R statistical software (“pwr” package) (14). Based on the results of a previous study that evaluates the impact of iso-inertial and gravitational resistance training protocols in professional athletes and evaluates the differences between the two protocols using iso-inertial tests (15), we estimate an effect size and a standard deviation of the data of 202 W and 153'97 W, respectively, for the primary outcome of our study (eccentric muscle power). No similar studies have been found to assess these differences with gravitational systems. Assuming an alpha risk of 0.05, a beta risk of 0.1 (power = 90%) and a dropout rate of 10% based on previous studies (16), the sample size required for this study is 30 participants (15 per group).

**Randomisation:** The randomization sequence will be created by the team's statistician and methodologist (DB) using the R statistical software (“blockrand” package) (14). This randomization will be performed by blocks of 4 and will be stratified by sex and age groups (60-66, 67-73, 74 or more) to balance groups in these variables. The randomization sequence will be sent to the administrative staff of the center responsible for recruitment of participants, who will match this sequence with the participants' membership numbers. In this way, the assignment will be hidden from the participants and research staff until the moment that the training program starts.

**Blinding:** Given the nature of the intervention, neither study participants nor the investigators monitoring the execution of the training protocols may be blinded from the assigned intervention. However, outcome assessors will not know the assignment of each participant.

**Data management:** Data will be stored and encoded in two Excel files to protect confidentiality before, during and after the study. The first of these files will be managed by the administrative staff of the center and will serve only to manage the recruitment of the participants and to match each of them with their intervention group. This file will contain the basic personal data of the participants: Name and surnames, age and sex. The second file will be managed entirely by the research team and will contain the registry of all the study variables in T0 and T1. This file will be entered and stored electronically in a password-protected Google Drive folder at the Universitat Internacional de Catalunya until the study is completed.

**Statistical methods:** For the statistical analysis we will use the R statistical software. Initially, we will present descriptive data of the subjects divided by sex: total numbers and percentages for qualitative variables, means and standard deviations for continuous quantitative variables, and medians and interquartile ranges for discrete quantitative variables.

For the main objective of the study, we will build a generalized linear regression model where the dependent variable will be the muscle power during the eccentric phase and the independent variables will be the group to which the patient has been assigned (control or intervention), some demographic variables (age and sex), and the degree of adherence to the training program (only for the experimental group).

For the secondary objectives of the study, we will build generalized linear regression models where the dependent variables will be each of the secondary outcomes and the independent variables will be the group to which the patient has been assigned (control or intervention), some demographic variables (age and sex), and the degree of adherence to the training program (only for the experimental group).

For all the previous models, we will check if they are valid by checking the different application assumptions (linearity, homocedasticity, normality and independence). Also, we will assess the inclusion of other variables as adjustment variables of the model. Following the variable selection process by Hosmer and Lemeshow (17), which begins by constructing univariate models for each of the independent variables, the final model will include those variables for which we observe a significant association with the dependent variable and which result in a better model. We will report the adjusted difference of means between groups along with its 95% confidence interval and the p-value of the comparison. The significance level will be 0.05. Also, we will conduct a gender subgroup analysis to explore the differences between men and women in the previous analyses.

Initially, these analyses will be performed for participants who complete the study ('complete-case analysis'). In case of dropouts or missing data, we will evaluate the volume and nature of these, explore data imputation strategies and, at least, report the best- and worst-case scenarios (18).

**Ethical considerations**

**Research ethics approval.** This protocol has submitted to the Drug Research Ethics Committee (CEIm) of the Universitat Internacional de Catalunya.

**Informed consent and materials.** The informed consent document can be obtained from investigators upon request.

**Confidentiality.** All information will be treated as strictly confidential, in accordance with the current regulations: Regulation (EU) 2016/679 of the European Parliament and of the Council of 27 April 2016 (GDPR) and Organic Law 3/2018 of 5 December on the Protection of Personal Data and Guarantee of Digital Rights. Participants will be identified by a code and only the research and work team with the right of access to the original data will be able to relate the data collected in the study to the identity of the participants. The identity of the participants will not be available to any other person except in the event of a medical emergency or legal requirement.

**Competing interests.** The investigators declare no conflicts of interest.

**Limitations.** This study has some limitations. Firstly, it will be developed in the context of only one sports center, which may hinder the achievement of the desired sample size. If the desired sample size is not reached, we will consider getting in touch with further sport centers where we could find potential participants.

Secondly, we did not include further physiological parameters that could provide more information on the functional and structural adaptations. If the study results confirm the hypotheses proposed, future research could include these parameters to further understand the underpinning physiological mechanisms behind the effects of iso-inertial and gravitational training.

Also, the impact of the proposed interventions will only be evaluated in the short-term. Future research could investigate whether the short-term changes observed are maintained in the long term.

Furthermore, we cannot be completely sure that, during the 6-week intervention time, participants do not do other strength work in the gym that may interfere with the intervention. If this happened, the study results could be affected. To minimise this potential bias, we explicitly instruct participants not to perform any strength work for the lower limbs while they are in the study. However, there might be cases were participants may not comply with our instructions.

Finally, regarding the assessment of outcomes related to muscle power, it is possible that the pre-intervention values will be slightly lower than the post-intervention ones due to the lack of familiarity of the participants with the exercises and the evaluation devices.

**Impact.** From a social and community point of view, the execution of this project would bring older adults into an active and dynamic setting like a gymnasium, which would also promote their social interaction with other users. Furthermore, this proposal encourages an inclusive intergenerational model in an environment where the young population is usually the protagonist. Previous studies have demonstrated the bidirectional benefits of the interaction between adults and young people and how this interaction fosters qualities such as empathy or solidarity in society.

Empowering older adults' confidence in their physical abilities can lead to changes in their physical and mental health. This project aims to focus on the benefits associated with physical health, not only in terms of analytical neuromuscular adaptations, but also on how these adaptations could be transferred to daily life activities. Most studies that evaluate the effect of strength training focus on gains in muscle strength or power. However, this project aims to go one step further, and explore what impact this type of training has on longevity markers such as the physical fitness or the risk of falls.

Furthermore, minimizing the longevity markers mentioned above would generate a decrease in the demand for health care and public health costs: medical visits, drug prescription, surgical interventions, or hospitalizations resulting from a lack of physical fitness.

In the field of physiotherapy, this project would generate new knowledge that would help clinical care practice and guide rehabilitation or prevention programs towards more effective exercises. Conducting the study in the gymnasium would also encourage multidisciplinary interaction between professionals (physiotherapists and fitness instructors) to achieve a more holistic approach to the needs of the older adults. It would also stimulate further research on eccentric strength work, whose benefits have been widely demonstrated in the young athletic population but barely so in older adults. The development of this project would help move away from a traditional generalist model in terms of exercise prescription and instead implement exercise programs tailored to the specific needs of the older adults, which would be a significant step forward in achieving better social health.

**Dissemination plans**. The results of the project will be published as a research article in a peer-reviewed, open-access journals that are indexed in the Journal of Citation Reports, such as “BMJ Open Sport & Exercise Medicine” or “BMC Geriatrics”.

In addition, we will attend and present the study findings in recognized conferences in this field of research, both nationally and internationally, such as the Congreso Nacional de Fisioterapia or the Geriatrics session of the [Global Conference on Physical Medicine and Rehabilitation](https://physical-medicine.magnusconferences.com/) (GCPR)., These results will also be communicated at research conferences aimed at promoting physical health in older adults or at conferences related to health science, sport science or community health.

Also, the study results will be discussed with the managers of the center where the intervention will be implemented. The objective of this is to help develop strength training programs that are tailored to older adults and that are based on the best scientific evidence. The research team will also contact other gymnasiums and rehabilitation centers to disseminate the study findings, as well as societies focused on older adults such as the Sociedad Española de Geriatría y Gerontología.

**Implementation**

1) **Work plan**

The proposed timeline and work plan for this project is the following:

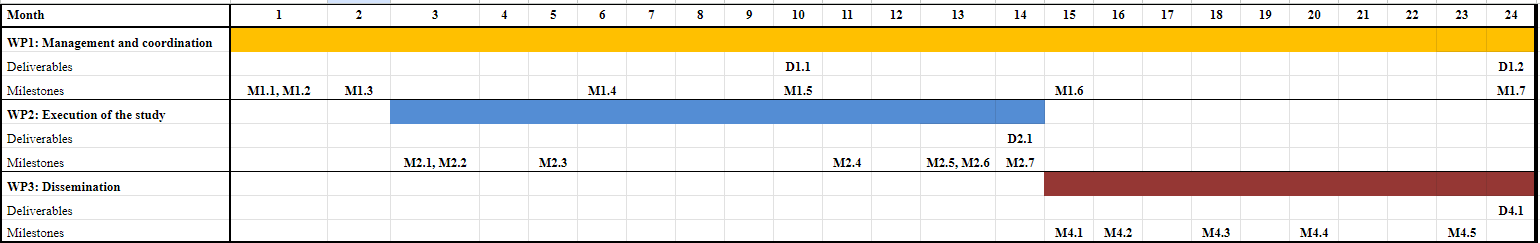
**Work Package 1: Management and Coordination**. The leader of this work package will be Dr. Aïda Cadellans Arróniz, and the other members will be Dr. Daniel Romero Rodríguez, Dr. Marc Madruga Parera, Dr. David Blanco.

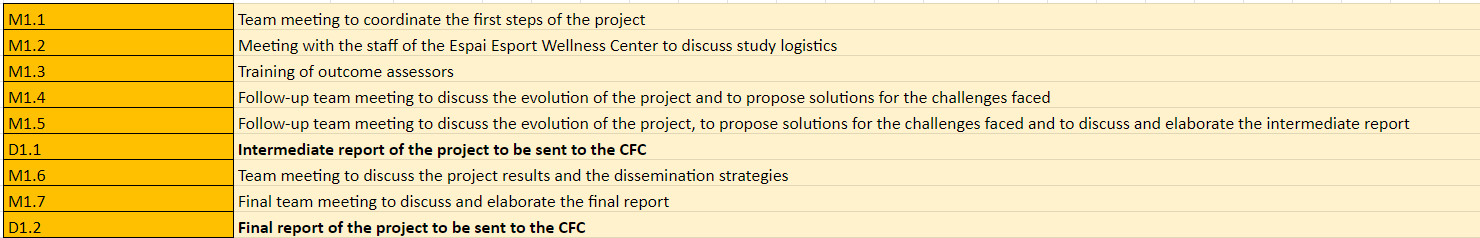
**Work Package 2: Study Execution**. The leader of this work package will be Dr. Aïda Cadellans Arróniz, and the other members will be Dr. Daniel Romero Rodríguez, Dr. Marc Madruga Parera, Dra. Ortega Cebrián, Flora Dantony and Dr. David Blanco.
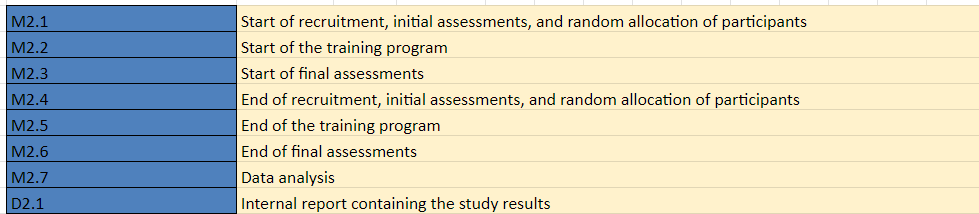
**Work Package 3: Study Dissemination**. The leader of this work package will be Dr. Aïda Cadellans Arróniz, and the other members will be Dr. Daniel Romero Rodríguez, Dr. Marc Madruga Parera, Dra. Ortega Cebrián, Flora Dantony and Dr. David Blanco.

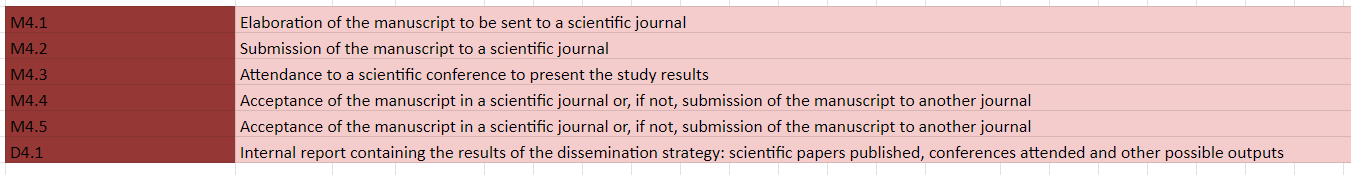

2) **Risk management plan:** The research team has identified the following risks:

| **Potential risks** | **Potential impact** | **Mitigation mechanism** |
| --- | --- | --- |
| Injuries of participants | Very low risk (as no adverse effects of resistance training in older adults have been reported) | If a participant gets injured while performing the tests or the training sessions, the potential physical damages that the participant may suffer will be covered by an insurance policy that the research team has purchased for this purpose. All adverse events and harms will be registered, analysed and reported together with the study results. |
| Recruitment problems | Medium risk | If reaching the desired sample size is not possible after the time specified in the work plan, we will consider getting in touch with further sport centers where we could find potential participants. |
| Compliance with the weekly program | Medium risk | If participants are unable to complete two sessions per week, they will be considered as compliant with the training program as long as they have completed a total of 12 sessions in 6 weeks, respecting the 48-hour break between sessions. |

**Cross-cutting aspects.** The proposed research is multidisciplinary as it is composed of four physiotherapists, one physical educator, and one research methodologist and statistician. The expertise of all these professionals guarantees that this project is solid and feasible.

Gender perspective has been considered in the composition of the research team, promoting an equal participation of women and men, and ensuring that there are no hierarchical gender relation patterns. In this regard, the Universitat Internacional de Catalunya has an Equality Unit, which ensures compliance with legislation on equal opportunities for men and women. Moreover, the gender perspective has been integrated into the different phases of the proposed research project: (a) the objectives and hypotheses of the study include the exploration of gender differences, (b) the recruitment will be stratified by sex to ensure equal distribution of men and women across the two study groups, (c) analyses will be first adjusted to gender and, second, performed by subgroups of men or women to explore potential differences.

This project aims to be socially inclusive by focusing on a population, the older adults, that is often overlooked in terms of public health interventions. Traditionally, society tends to consider the elderly as vulnerable in order to protect their health. Older adults are often isolated in their own group and are then removed from a young and active society, which increases their fragility and limits their capacities. Increasing the physical activity of the older adults by focusing on their possibilities and not so much on their limitations, as it is often done in a rehabilitative or geriatric context, is key to improving their health and quality of life.

The research results will be published in open access. To ensure that, there are funds at the Department of Physiotherapy of the Universitat Internacional de Catalunya available that are dedicated specifically to cover open access taxes. Once the study is published, a file containing all coded data will be shared on the repository CORA (Repositori de dades de Recerca) to meet the FAIR data principles (Findable, Accessible, Interoperable and Reusable). This file will be freely accessible to all scientific communities under the license Creative Commons Attribution NonCommercial NoDerivs (CC-BY-NC-ND).

**References.**

7. Maroto-Izquierdo S, García-López D, Fernandez-Gonzalo R, Moreira OC, González-Gallego J, de Paz JA. Skeletal muscle functional and structural adaptations after eccentric overload flywheel resistance training: a systematic review and meta-analysis. J Sci Med Sport

8. Tesch PA, Fernandez-Gonzalo R, Lundberg TR. Clinical applications of iso-inertial, eccentric-overload (YoYoTM) resistance exercise. Front Physiol. 2017;8(APR).

15. Gual, G.; Fort-Vanmeerhaeghe, A.; Romero-Rodríguez, D.; Tesch, P.; Costa L. Eccentric overload by inertial resistance training: Effects on patellar tendinopathy prevention and muscle power in jumping sports. Congr Eur Coll Sport Sci. 2013;

16. Madruga-Parera M, Bishop C, Fort-Vanmeerhaeghe A, Beato M, Gonzalo-Skok O, Romero-Rodríguez D. Effects of 8 Weeks of Isoinertial vs. Cable-Resistance Training on Motor Skills Performance and Interlimb Asymmetries. J Strength Cond Res. 2022 May 1;36(5):1200-1208.

17. Hosmer DW. L. Strategies and Methods for Logistic Regression. In Applied Logistic Regression. Hoboken, NJ, USA, 2000; pp 91–142.

18. Jakobsen, J.C., Gluud, C., Wetterslev J et al. When and how should multiple imputation be used for handling missing data in randomised clinical trials – a practical guide with flowcharts. BMC Med Res Methodol. 2017;17(162).

**Annex 1**

6-week training program (2 sessions per week). *These variations will be performed following markings on the floor, which will indicate the depth and direction of the frontal and side lunges.

|  | Week | | | | | |
| --- | --- | --- | --- | --- | --- | --- |
|  | 1 | 2 | 3 | 4 | 5 | 6 |
| **Exercise 1** | Front lunge | Front lunge | Front lunge | Same exercises of week 3 with variations of directions and distances* | Front lunge | Same exercises of week 5 with variations of directions and distances* |
| **Exercise 2** | Side lunge | Side lunge | Side lunge |  | Side lunge |  |
| **Exercise 3** | Front lunge with hand grip | Front lunge with hand grip | Front lunge with hand grip |  | Front lunge with hand grip |  |
| **Series** | 2 | 2 | 3 | 3 | 3 | 3 |
| **Repetitions** | between 6 and 8 | between 6 and 8 | 10 | 10 | 10 | 10 |
| **RPE** | between 3 and 5 | between 3 and 5 | between 3 and 5 | between 3 and 5 | between 6 and 8 | between 6 and 8 |

**Annex 2**

*Images of the two groups of exercises.*

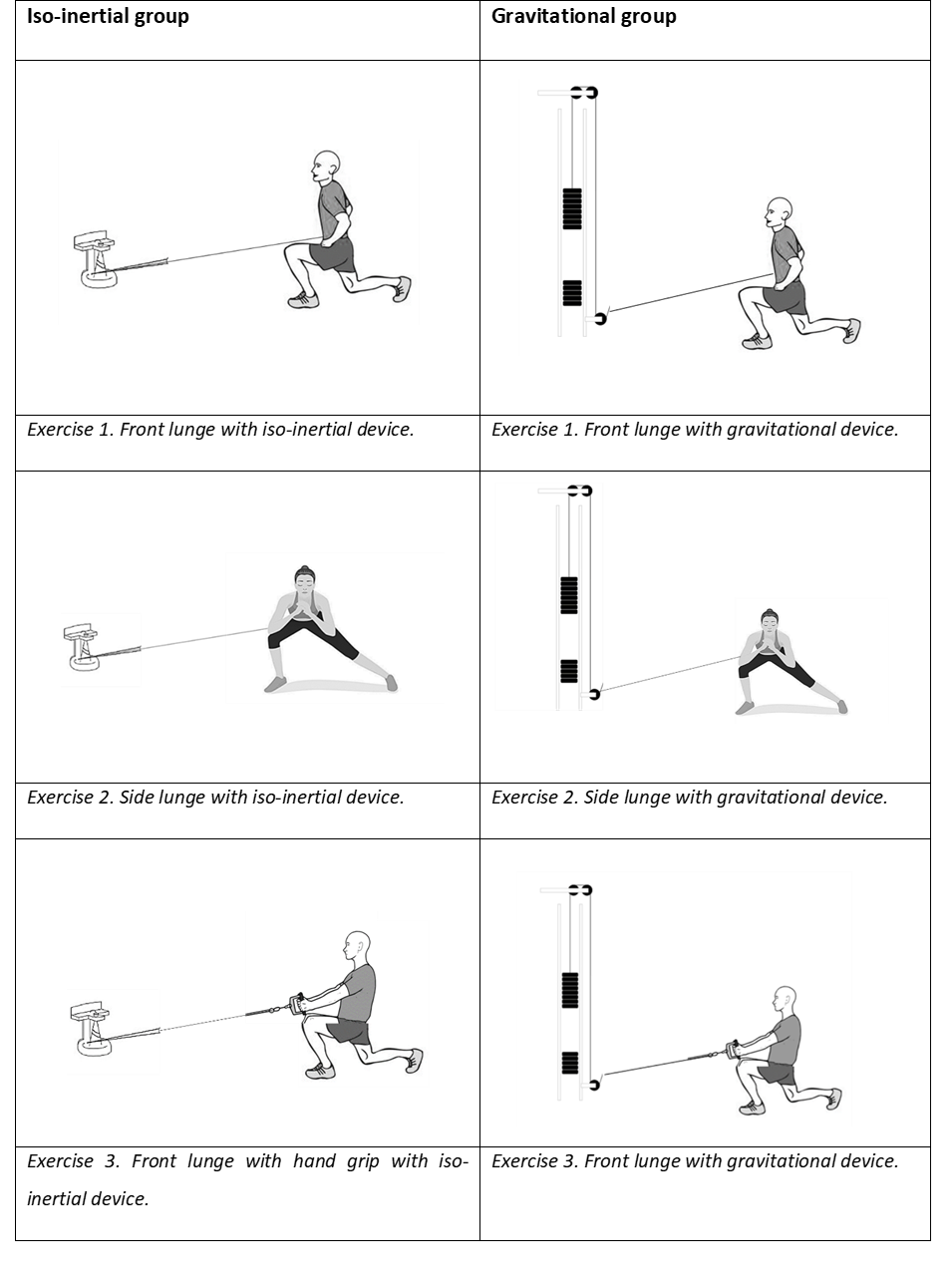
